# Forward Stroke Volume and Ejection Fraction Estimation in Mitral Regurgitation: A Computational Hemodynamic Approach

**DOI:** 10.1101/2025.08.27.25334572

**Authors:** Amirahmad Saeidi, Mobina Ghajar, Sara Tarkiani, Amin Ramezani

## Abstract

We developed and validated a closed loop lumped parameter cardiovascular model incorporating a regurgitant pathway across the mitral valve and left ventricular outflow tract dynamics to simulate mitral regurgitation (MR) and quantify its effects on forward stroke volume and forward ejection fraction. MR is a common valvular disorder in which preserved ejection fraction may mask a significant reduction in forward flow. Validated across a spectrum of MR severities, the model consistently demonstrated that, despite preserved total stroke volume and conventional EF, both forward stroke volume and forward ejection fraction declined markedly as regurgitation increased. These results accurately replicate the clinical paradox of MR, where standard EF metrics can conceal hemodynamic compromise. By providing direct quantification of forward output, this framework offers a physiologically faithful tool for assessing true hemodynamic burden in MR. It underscores the limitations of traditional EF based assessment and highlights LVOT based flow metrics as more reliable indicators for clinical evaluation and treatment planning in MR.

## 1 Introduction

Mitral regurgitation (MR), also referred to as mitral insufficiency, is one of the most common valvular heart diseases worldwide. It occurs when the mitral valve fails to close properly during systole, allowing blood to leak backward from the left ventricle (LV) into the left atrium (LA). The condition may arise due to primary causes—such as mitral valve prolapse, myxomatous degeneration, or rheumatic disease—or secondary causes related to left ventricular remodeling and dysfunction, often seen in ischemic or dilated cardiomyopathy. The disease burden is particularly notable in older adults; epidemiological studies estimate that moderate to severe MR affects approximately 10% of individuals over the age of 75. As life expectancy increases globally, the prevalence and clinical relevance of MR are expected to rise significantly [1–3].

From a pathophysiological perspective, MR imposes a chronic volume overload on the left side of the heart. With each heartbeat, a portion of the stroke volume regurgitates into the left atrium instead of progressing into the systemic circulation. Importantly, although total stroke volume may appear preserved or even increased, the forward stroke volume-which determines effective systemic perfusion-is often significantly reduced. This mismatch can lead to underestimation of disease severity, especially in standard echocardiographic evaluations. This retrograde flow leads to left atrial enlargement, elevated pulmonary pressures, and progressive left ventricular dilation and dysfunction. Over time, these changes can trigger a cascade of complications including atrial fibrillation, pulmonary hypertension, right ventricular failure, and eventually, overt congestive heart failure. Despite this progressive nature, many patients remain asymptomatic for years, which highlights the importance of timely recognition and monitoring of the disease’s course [4–7]. Clinically, the presentation of MR is diverse and heavily influenced by the severity and chronicity of regurgitation. In mild cases, patients may remain entirely asymptomatic and the diagnosis is often made incidentally during routine physical examination or echocardiography. As the condition advances, common symptoms include exertional dyspnea, fatigue, orthopnea, and reduced exercise tolerance-symptoms that overlap with other forms of heart failure, making clinical distinction challenging. In acute MR, often caused by papillary muscle rupture post-myocardial infarction or infective endocarditis, patients may present with sudden pulmonary edema and cardiogenic shock, requiring urgent surgical intervention [8, 9].

Other cardiac abnormalities may also lead to alterations in stroke volume dynamics and should be considered in the differential diagnosis of mitral regurgitation. For instance, heart failure with preserved ejection fraction (HFpEF) can present with normal or near-normal EF but reduced forward stroke volume due to impaired ventricular filling. Similarly, significant aortic regurgitation leads to increased total stroke volume while forward output is compromised by retrograde flow into the left ventricle, mimicking the hemodynamic profile seen in MR. Cardiomyopathies, such as dilated or hypertrophic types, can cause both systolic and diastolic dysfunction, altering ventricular compliance and reducing effective output. Conduction abnormalities, including left bundle branch block (LBBB), may cause mechanical dyssynchrony that affects the timing and coordination of ventricular contraction, leading to inefficient ejection. In such scenarios, standard metrics like EF may be misleading, reinforcing the need for comprehensive forward flow assessment. Differentiating MR from these conditions requires careful hemodynamic analysis, especially when forward stroke volume is disproportionately low relative to total stroke volume [10].

The accurate assessment of MR severity and its impact on left ventricular function is crucial in determining optimal timing for surgical or transcatheter intervention. Current diagnostic methods include transthoracic and transesophageal echocardiography, cardiac MRI, in some cases, cardiac catheterization. Parameters such as effective regurgitant orifice area (EROA), regurgitant volume, and LV ejection fraction (EF) are commonly used. However, these parameters can be affected by loading conditions and operator variability, potentially leading to inconsistent decision-making in borderline cases. Moreover, the hemodynamic burden of MR is not fully captured by conventional measurements, for instance a preserved EF might mask underlying LV dysfunction due to the lowered afterload from regurgitant flow. Therefore, more nuanced assessment methods-particularly those integrating dynamic and patient-specific cardiac mechanics-are needed to improve diagnostic precision [11, 12]. Computational modeling offers a promising avenue for capturing the complex hemodynamics of MR beyond conventional metrics, providing deeper insight into disease progression and ventricular workload under varying pathological conditions.

Despite advances in diagnostic imaging, accurate quantification of effective cardiac output in patients with mitral regurgitation (MR) remains a clinical challenge. Traditional parameters such as left ventricular ejection fraction (LVEF) often fail to reflect true hemodynamic compromise because they do not account for regurgitant volume. This can lead to misclassification of disease severity and delays in appropriate intervention. There is a clear need for alternative metrics that better capture the forward flow component of cardiac output to improve risk stratification and clinical decision-making. This study aims to address this gap by developing and validating a computational framework for estimating forward stroke volume (FSV) and ejection fraction in MR using a lumped-parameter model informed by clinically relevant flow dynamics.

The remainder of this paper is organized as follows. Section II presents the modeling framework, including the lumped-parameter cardiovascular model, governing equations, and the method for simulating MR and calculating forward flow. Section III reports key simulation results, including comparisons across different severities of MR Section IV discusses these findings in light of clinical relevance and existing diagnostic limitations. Section V concludes with a summary of the main insights from this study.

## 2 Methodology

### 2.1 Physiological Modeling of the Cardiovascular System

In this study, we use a closed-loop lumped-parameter model (LPM) to simulate how the cardiovascular system works. This model is based on the framework introduced in paper [13–15]. It uses electrical circuit-like components to represent how blood moves and how pressure and volume change in different parts of the heart and blood vessels.

Each chamber of the heart—the left and right atria (LA and RA) and the left and right ventricles (LV and RV)—is modeled using a time-varying stiffness (called elastance), which helps calculate pressure based on the volume of blood in the chamber at any time. The blood vessels in the lungs and the body are modeled using elements that represent resistance, flexibility (compliance), and inertia, similar to how they behave in real life. Heart valves are modeled as one-way switches (like electrical diodes) that allow blood to flow in only one direction, while also considering some energy loss as blood flows through them.

### 2.2 Governing Equations for Volume and Pressure Dynamics

The model equations are based on basic physical laws that describe how mass and movement are conserved [16]. In the heart, the amount of blood in each chamber changes depending on how much blood flows in and out.

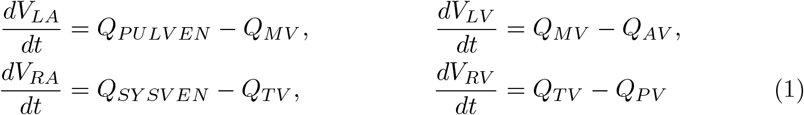

where:

*V*_*LA*_, *V*_*LV*_, *V*_*RA*_, and *V*_*RV*_ are the blood volumes in the respective chambers.

*Q*_*PULVEN*_ and *Q*_*SYSVEN*_ are venous inflows from the pulmonary and systemic veins.

*Q*_*MV*_ and *Q*_*TV*_ are flows through the mitral and tricuspid valves.

*Q*_*AV*_ and *Q*_*PV*_ are flows through the aortic and pulmonary valves.

Changes in arterial and venous pressures are expressed using compliance relations:

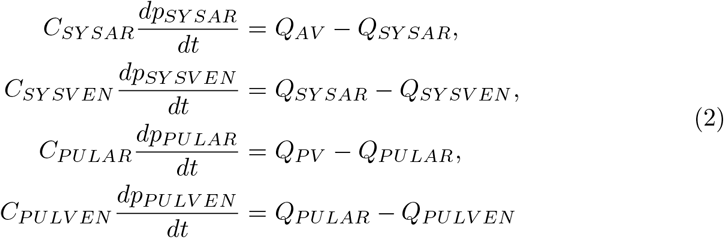

where:

*p*_*SYSAR*_, *p*_*SYSVEN*_, *p*_*PULAR*_, and *p*_*PULVEN*_ are pressures in arteries and veins.

*C* is the compliance (inverse of stiffness) of the vessel segments.

Blood inertia is modeled using inductive elements:

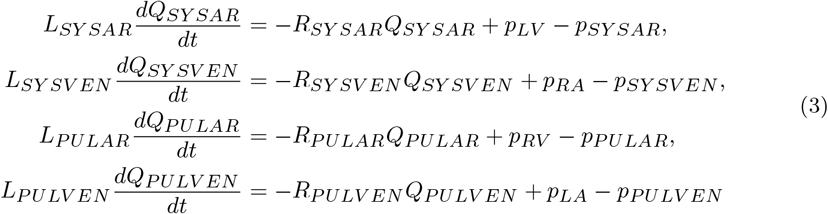

where:

*Q*_*SYSAR*_, *Q*_*SYSVEN*_, *Q*_*PULAR*_, and *Q*_*PULVEN*_ are flow rates through vessels.

*R* is the resistance to flow, and *L* is the inertance.

*p*_*LA*_, *p*_*RA*_, *p*_*LV*_, and *p*_*RV*_ are pressures in the left/right atria and ventricles. The governing equations for blood volume and pressure dynamics presented here are adopted directly from the models in [13].

### 2.3 Pressure and Valve Flow Definitions

Chamber pressures are defined using the time-varying elastance model based on [13]:

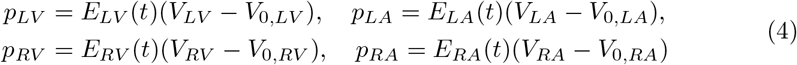

Valve flow rates depend on the pressure gradient and valve resistance:

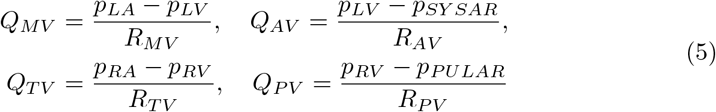

where:

*R*_*MV*_ is the mitral valve resistance, *R*_*AV*_ is the aortic valve resistance, *R*_*TV*_ is the tricuspid valve resistance, and *R*_*PV*_ is the pulmonary valve resistance.

Valves operate as non-ideal diodes:

- If *p*_in_ *> p*_out_, *R ≈ R*_min_
- If *p*_in_ *≤ p*_out_, *R ≈ R*_max_

Mitral valve leakage, also known as mitral regurgitation, occurs when the valve between the left atrium and left ventricle does not close completely.

As a result, some blood flows backward into the left atrium during ventricular contraction.

This backflow is represented in the model using a leakage resistance parameter, *R*_*m_leak*_.

### 2.4 Time-Varying Elastance

Elastance is a measure of the stiffness of a cardiac chamber [17]. It tells us how much the pressure increases when a certain amount of blood volume is added to the chamber. In other words, elastance describes how responsive a chamber is in generating pressure for a given change in volume—higher elastance indicates greater stiffness and stronger contractility.

To capture the periodic contractile behavior of each cardiac chamber during the cardiac cycle, we define an activation function *ϕ*(*t*) using a piecewise cosine waveform. This function determines the timing and magnitude of the chamber’s active contraction and relaxation phases.

The elastance of each chamber is then modeled as a combination of passive and active components. The passive part represents the baseline stiffness of the chamber, while the active part varies over time according to the activation function. This allows us to simulate how the stiffness of the chamber increases during systole (contraction) and decreases during diastole (relaxation), reproducing the dynamic behavior of the heart throughout each heartbeat.

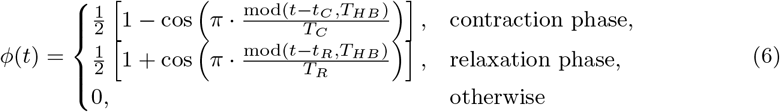

where *t*_*C*_ and *t*_*R*_ denote the onset times of contraction and relaxation, respectively; *T*_*C*_ and *T*_*R*_ represent the durations of contraction and relaxation; and *T*_*HB*_ is the heartbeat period. Note that *t*_*R*_ = *t*_*C*_ + *T*_*C*_. The chamber elastance is:

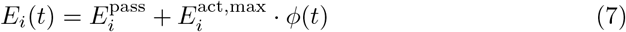

The time-varying elastance formulation is taken without modification from [13] to model dynamic chamber stiffness during the cardiac cycle.

### 2.5 Model Validation in Healthy Conditions

To ensure the accuracy of our lumped-parameter cardiovascular model, we validated it under normal physiological conditions. The model was simulated with a heart rate of 60 beats per minute, and the outputs were compared to established clinical benchmarks.

Key simulation results include:

- End-Diastolic Volume (EDV): 120.00 mL
- End-Systolic Volume (ESV): 43.14 mL
- Stroke Volume (SV): 76.86 mL
- Cardiac Output (CO): 4611.68 mL/min
- Ejection Fraction (EF): 64.05%
- Peak Systolic Pressure (LV): 127.78 mmHg

In addition to these standard measurements, we also validated the model using a forward-flow–based approach. This method estimates forward stroke volume (FSV) by integrating the velocity of blood across the left ventricular outflow tract (LVOT), a technique used in clinical echocardiography. From this simulation, we obtained:

- LVOT Velocity-Time Integral (VTI): 23.70 cm
- Forward Stroke Volume (FSV): 74.47 mL
- Forward Ejection Fraction (FEF): 62.98%

The Pressure–Volume (PV) loop for the left ventricle (LV), shown in Fig. 2, illustrates the characteristic rectangular cycle of cardiac contraction and relaxation. The loop captures the full cardiac cycle, including isovolumic contraction, ejection, isovolumic relaxation, and filling phases. The loop shape, peak pressures, and stroke volume are all consistent with expected healthy heart dynamics.

**Fig. 1.**
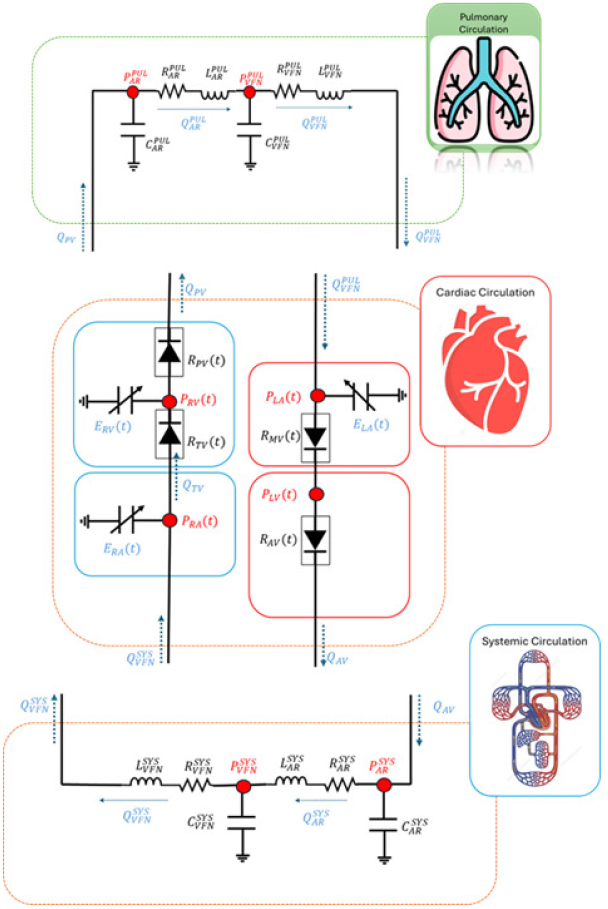
Lumped-parameter model of the closed-loop cardiovascular system.

**Fig. 2.**
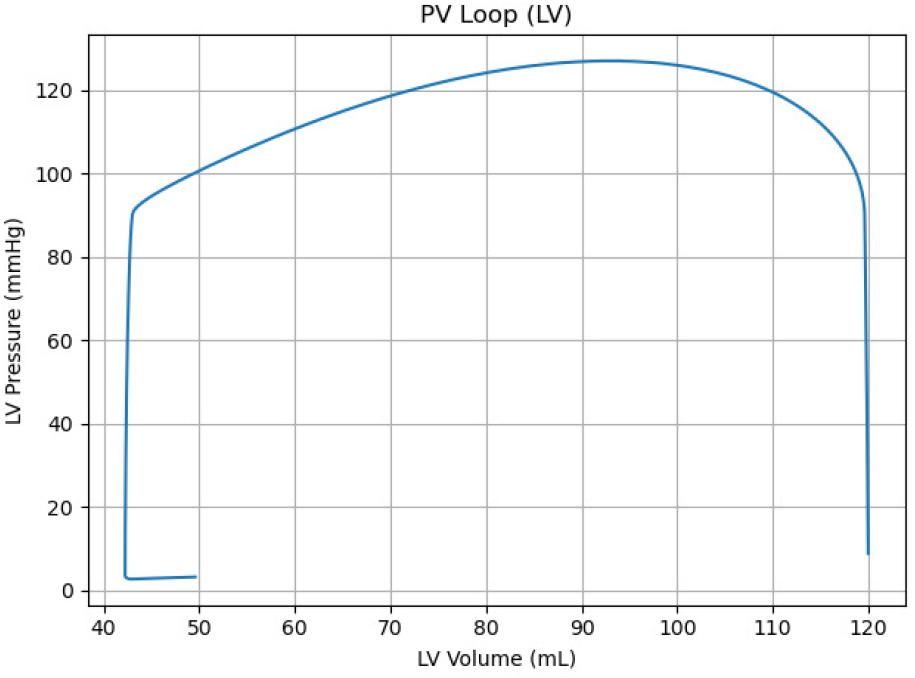
Pressure–Volume (PV) loop of the left ventricle under healthy conditions.

These results confirm that the model is able to reproduce realistic cardiovascular dynamics under baseline healthy conditions, providing a reliable foundation for simulating pathological cases in later sections.

### 2.6 Mitral Valve Regurgitation and Forward Flow Calculation

To simulate mitral valve regurgitation, we introduced a leakage pathway into the lumped-parameter cardiovascular model by adding a backward flow resistance parameter *R*_m___leak_ across the mitral valve. Lower values of this resistance correspond to more severe regurgitation, allowing more blood to flow backward from the left ventricle into the left atrium during systole. By varying *R*_m___leak_, we model different severities of mitral insufficiency and analyze their impact on forward cardiac output.

In mitral regurgitation, the stroke volume measured by traditional end-diastolic and end-systolic volumes may significantly overestimate the effective systemic output, as part of the blood is regurgitated into the left atrium. Thus, forward stroke volume (FSV), calculated via the left ventricular outflow tract (LVOT) velocity-time integral (VTI), provides a more clinically relevant index of cardiac efficiency in such pathologies. This approach better reflects the actual volume of blood effectively ejected into the aorta, especially in the presence of regurgitant lesions. In our model, we simulate mitral regurgitation by introducing a regurgitant pathway across the mitral valve, allowing us to study its impact on forward cardiac output. The methodology follows the clinical principles described in [18],a technique used in clinical echocardiography [7], where LVOT VTI is used to quantify forward ejection volume.

To calculate FSV, we first estimate the aortic valve flow *Q*_*a*_(*t*) :

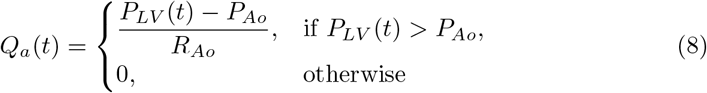

Here, *P*_*LV*_ (*t*) is the left ventricular pressure computed from the time-varying elastance model, *P*_*Ao*_ is the assumed aortic pressure (e.g., 90 mmHg) [19], and *R*_*Ao*_ is the resistance across the aortic valve.

The instantaneous blood velocity through the LVOT is then given by:

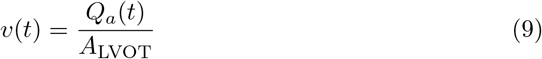

where *A*_LVOT_ is the cross-sectional area of the LVOT.

The LVOT velocity-time integral (VTI) is obtained by integrating the velocity over the systolic phase:

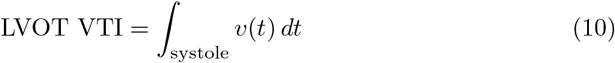

Finally, the forward stroke volume is computed as:

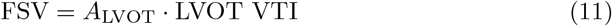

To compute these values, we used standard anatomical assumptions from echocardiography. The LVOT area was derived using the circular area formula, assuming an LVOT diameter *d* of 2.0 cm:

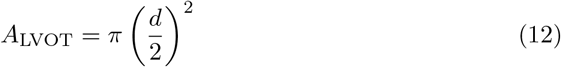

Additionally, the left ventricular end-diastolic volume (LVEDV) was estimated using the Teichholz formula [20] based on the end-diastolic diameter (LVEDD):

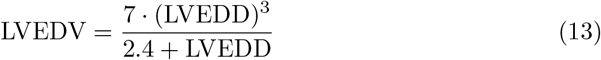

This formulation allows for quantification of the true forward output into the systemic circulation, which is especially important when mitral regurgitation causes a portion of the stroke volume to be lost to backflow. To estimate this, we model the left ventricle as a prolate ellipsoid—a clinically accepted approximation widely used in echocardiography when direct volumetric imaging is unavailable. In our simulations, we assumed an LVOT diameter of 2.0 cm and an LVEDD of 5.0 cm, resulting in an estimated LVOT area (*A*_LVOT_) of approximately 3.14 cm^2^ and an end-diastolic volume (LVEDV) of approximately 118.24 mL.

### 2.7 Simulation of Cardiac Abnormality

The model also enables the simulation of pathological conditions by adjusting specific physiological parameters. In this section, we explore common cardiac abnormality, which is mitral valve regurgitation.

*Mitral Valve Regurgitation:* To represent a faulty mitral valve that does not close completely, we introduce a regurgitation effect by modifying the parameter 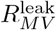 which controls the resistance to backward flow. In a healthy heart, the mitral valve prevents blood from flowing back into the left atrium during ventricular contraction. However, in the presence of regurgitation, some blood leaks back into the atrium. This is modeled by reducing 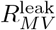 to allow partial reverse flow from the left ventricle to the left atrium during systole. This setup enables the study of how mitral leakage affects intrachamber pressures, flow patterns, and volume dynamics.

### 2.8 Model Parameters

Tables 1a, 1b, 1c, and 1d summarize the key physiological parameters used in our simulations, including timing parameters, valve characteristics, ventricular properties, and circulatory system parameters, which collectively define the baseline conditions for the modeled cardiovascular system. The mitral regurgitation parameter is also set to *R*_M,leak_ = 0.01–10 (*mmHgs/mL*).

**Table 1.**
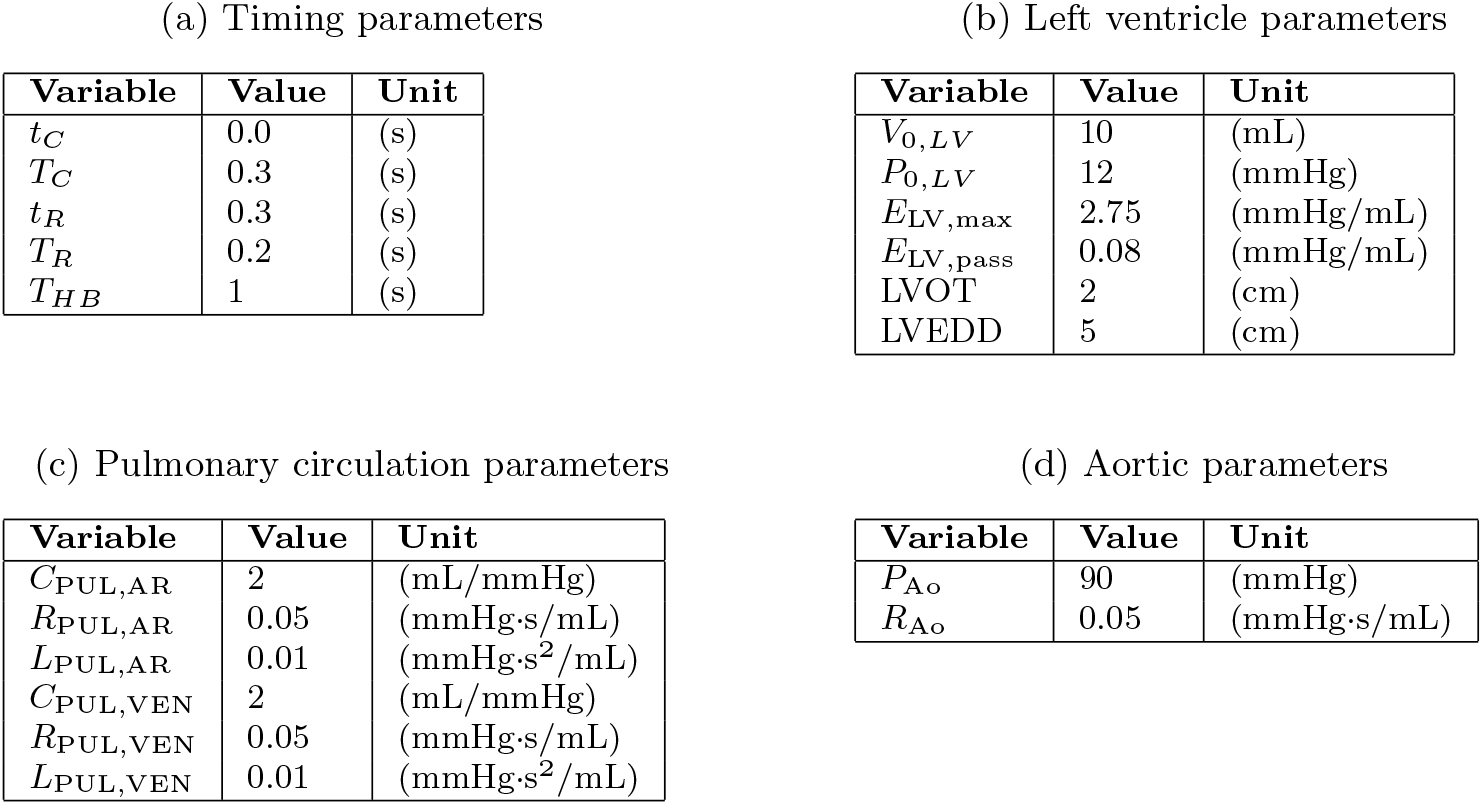
Model parameters.

| Variable | Value | Unit |
| --- | --- | --- |
| $t_C$ | 0.0 | (s) |
| $T_C$ | 0.3 | (s) |
| $t_R$ | 0.3 | (s) |
| $T_R$ | 0.2 | (s) |
| $T_{HB}$ | 1 | (s) |

(b) Left ventricle parameters
| Variable | Value | Unit |
| --- | --- | --- |
| $V_{0,LV}$ | 10 | (mL) |
| $P_{0,LV}$ | 12 | (mmHg) |
| $E_{LV,max}$ | 2.75 | (mmHg/mL) |
| $E_{LV,pass}$ | 0.08 | (mmHg/mL) |
| LVOT | 2 | (cm) |
| LVEDD | 5 | (cm) |

(c) Pulmonary circulation parameters
| Variable | Value | Unit |
| --- | --- | --- |
| $C_{PUL,AR}$ | 2 | (mL/mmHg) |
| $R_{PUL,AR}$ | 0.05 | (mmHg·s/mL) |
| $L_{PUL,AR}$ | 0.01 | (mmHg·s<sup>2</sup>/mL) |
| $C_{PUL,VEN}$ | 2 | (mL/mmHg) |
| $R_{PUL,VEN}$ | 0.05 | (mmHg·s/mL) |
| $L_{PUL,VEN}$ | 0.01 | (mmHg·s<sup>2</sup>/mL) |

**Table 1:** Model parameters
| Variable | Value | Unit |
| --- | --- | --- |
| $P_{Ao}$ | 90 | (mmHg) |
| $R_{Ao}$ | 0.05 | (mmHg·s/mL) |

## 3 Results

### 3.1 Normal Heart Simulation

To establish a baseline, we simulated the cardiovascular system under normal physiological conditions with a heart rate of 60 beats per minute. Fig. 3 shows the left ventricular pressure–volume (PV) loop for a single heartbeat, demonstrating the characteristic four-phase cycle of the cardiac cycle: isovolumic contraction, ejection, isovolumic relaxation, and filling.

**Fig. 3.**
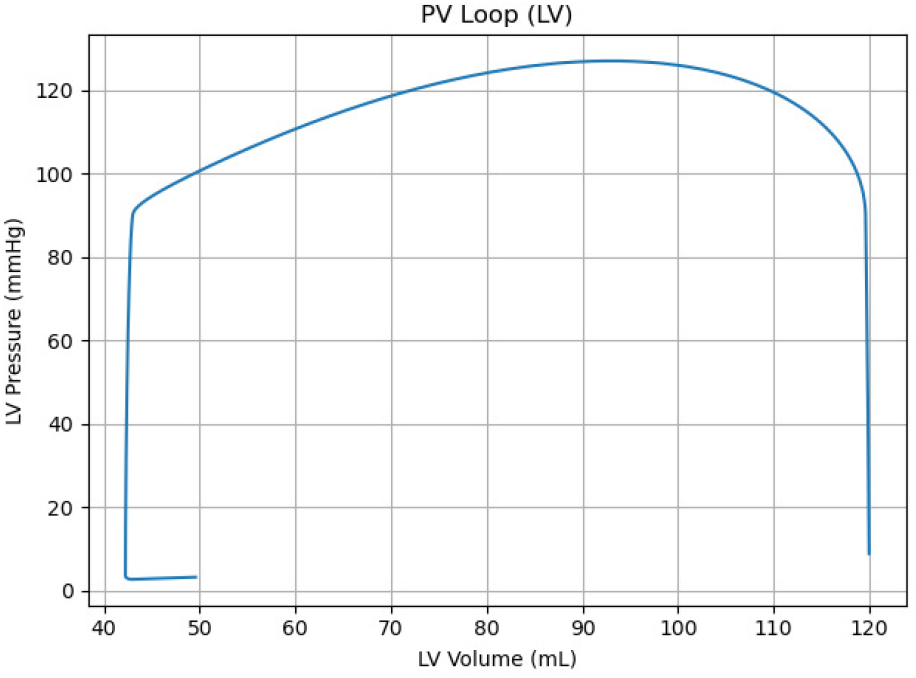
PV loop of the left ventricle under normal conditions.

Fig. 4 extends the simulation to a 10-second duration, capturing multiple cardiac cycles. The PV loop remains consistent over time, confirming stable cardiac performance under normal conditions.

**Fig. 4.**
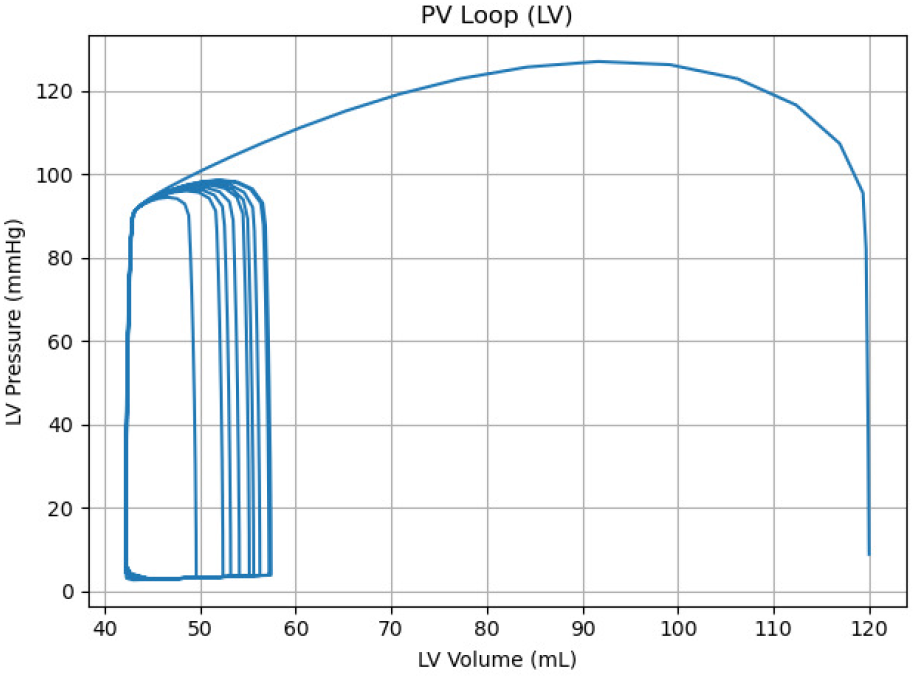
Left ventricular PV loops over a 10-second simulation.

Fig. 5 and 6 show the time-varying pressure and volume of the left ventricle (LV), respectively, throughout the simulation. These waveforms accurately reflect the physiological behavior of a healthy LV, including the timing of systolic contraction and diastolic filling.

**Fig. 5.**
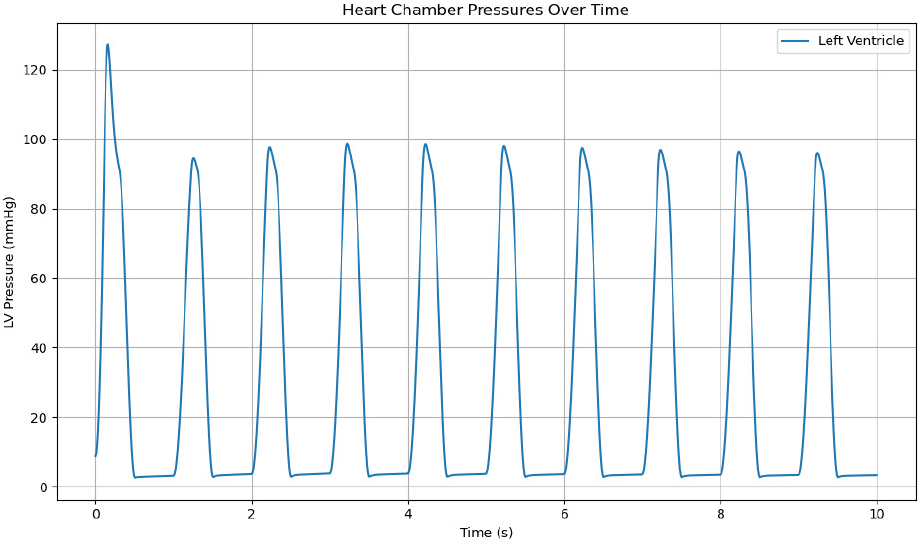
Left ventricular pressure over time during normal cardiac cycles.

**Fig. 6.**
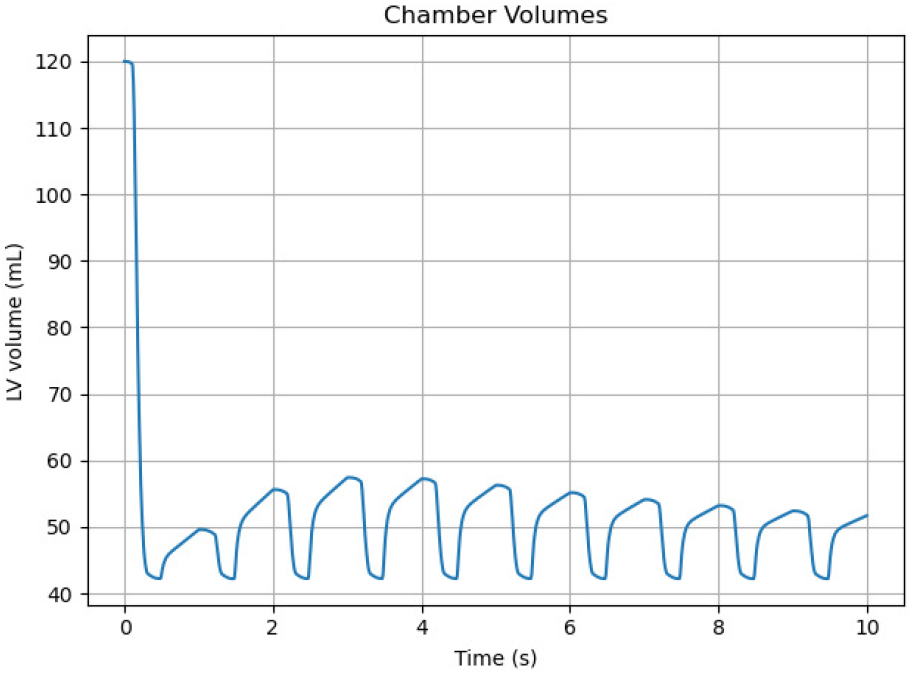
Left ventricular volume over time during normal cardiac cycles.

### 3.2 Effect of Mitral Valve Leakage

To examine the impact of mitral valve regurgitation, we simulated different levels of leakage by varying the resistance to backward flow through the mitral valve, denoted as *R*_m___leak_. Lower values of *R*_m___leak_ correspond to more severe regurgitation.

Fig. 7 shows the pressure–volume (PV) loops for the left ventricle under seven different values of *R*_m___leak_, ranging from 0.01 to 10.

**Fig. 7.**
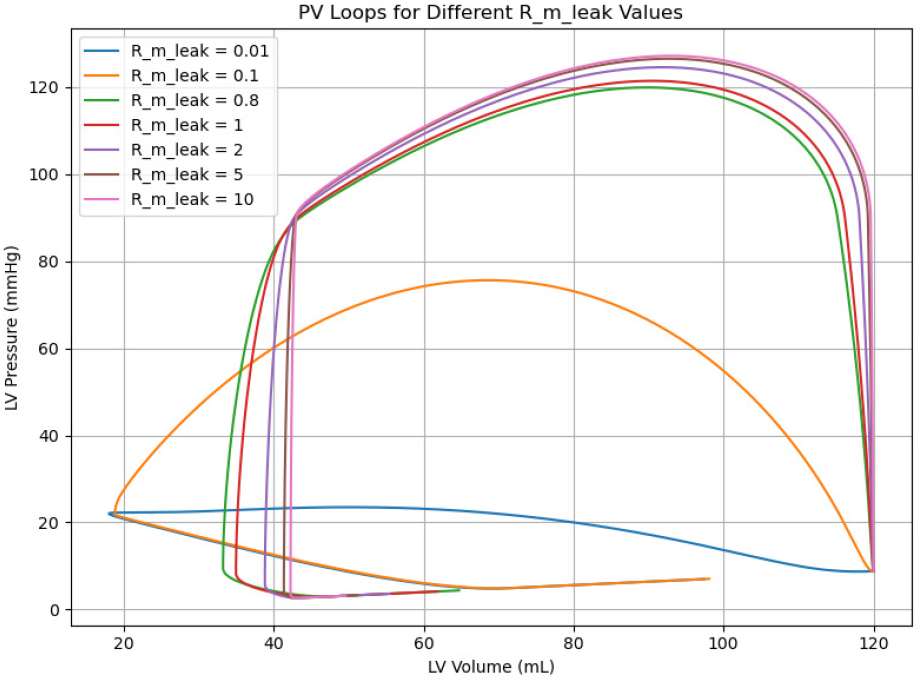
Left ventricular PV loops for varying *R*_m___leak_ values, representing different levels of mitral valve leakage.

The corresponding hemodynamic parameters are summarized in Table 2.

**Table 2.** Effect of Mitral Regurgitation (*R*_m___leak_) on Forward Hemodynamic Metrics.

| $R_{m\_leak}$ | LVOT VTI (cm) | FSV (mL) | LVEDV (mL) | FEF (%) |
| --- | --- | --- | --- | --- |
| 0.01 | 0.000 | 0.000 | 118.24 | 0.00 |
| 0.10 | 0.000 | 0.000 | 118.24 | 0.00 |
| 0.80 | 16.36 | 51.38 | 118.24 | 43.46 |
| 1.00 | 17.75 | 55.76 | 118.24 | 47.16 |
| 2.00 | 20.86 | 65.54 | 118.24 | 55.43 |
| 5.00 | 22.97 | 72.15 | 118.24 | 61.02 |
| 10.00 | 23.71 | 74.47 | 118.24 | 62.98 |

## 4 Discussion

As a highly prevalent valvular disorder, Mitral regurgitation is defined by incomplete mitral valve closure during systole leading to the retrograde flow of blood into the left atrium from the left ventricle. This condition is mostly subsequent to mitral valve prolapse which generally is attributable to myxomatous degeneration. Other plausible causes include rheumatic heart disease, infective endocarditis, papillary muscle rupture, degenerative valve disease and congenital abnormalities such as cleft mitral valve. MR can also occur secondary to underlying conditions such as left ventricle dilatation, ischemic cardiomyopathy, and atrial fibrillation [1, 2]. This common condition could eventually result in increased preload, reduced afterload, elevated ejection fraction, and a supranormal total stroke volume. However, forward stroke volume is diminished as a consequence of regurgitation. MR severity is further classified based on regurgitant fraction, such that values over 50% indicate severe MR, 30 *−* 50% suggests moderate MR, and values below 30% reflect mild MR [21]. Our model-based findings corroborate the theoretical framework that forward stroke volume is a more reliable indicator of hemodynamic function than LVEF in moderate to severe MR [18, 22]. Acute MR, such as that caused by papillary muscle rupture, imposes a sudden increase in preload. In the absence of adequate compensatory mechanisms, left atrial pressure increases markedly, giving rise to rapid onset of pulmonary congestion and edema. Despite an apparently preserved or elevated LVEF, forward stroke volume is significantly diminished, necessitating immediate clinical intervention [23]. Over time, chronic MR induces a range of adaptive remodeling processes in the left ventricle to accommodate the ongoing volume overload. These changes include left ventricular dilation with eccentric hypertrophy, increased diastolic compliance to lower wall tension, and progressive left atrial dilation [24]. It is important to note that, beyond MR, other abnormalities such as heart failure with preserved ejection fraction (HFpEF), cardiomyopathies, aortic regurgitation and left bundle branch block (LBBB) may present with altered cardiac dynamics [25–29]. Similar to MR, EF is an unreliable indicator of hemodynamic status in HFpEF, often remaining within the normal range despite significant reduction in forward stroke volume and cardiac output, due to several abnormalities including impaired ventricular filling, systolic and diastolic dysfunction, ventricular-atrial stiffening, and abnormal chronotropic and autonomic regulation. This limitation arises from EF’s nature as a ratio of stroke volume to enddiastolic volume, allowing it to remain normal even when both absolute volumes are markedly reduced. [25]. Notably, hypertrophic and restrictive cardiomyopathies may mimic HFpEF, presenting with a preserved EF masking a compromised cardiac output which could be attributable to significant diastolic dysfunction [28, 29]. Aortic regurgitation (AR) is a form of valvular heart disease characterized by improper closure of the aortic valve leading to retrograde blood flow from the aorta into the left ventricle [26]. In AR, as in MR, effective forward output may decline despite an elevated total stroke volume when compensatory mechanisms, such as increased end-diastolic volume and left ventricle dilation are insufficient, as seen in acute AR, potentially resulting in pulmonary edema, hypotension and cardiogenic shock [30]. Left bundle branch block (LBBB) is an intraventricular conduction disturbance that impairs the normal sequence of ventricular activation. This electrical dyssynchrony diminishes contractile efficiency and increases energy expenditure, ultimately contributing to left ventricular dysfunction and a reduction in EF [27, 31, 32]. Notably, recent evolution of computational modeling techniques has significantly enhanced the assessment of hemodynamics in conditions such as MR, yielding superior accuracy compared to conventional metrics alone. Such advancements contribute to a more precise characterization of cardiac performance and ventricular stress, enabling more prompt and evidence-based clinical interventions [33, 34].

On the whole, meticulous diagnostic evaluation is essential when assessing the described abnormalities, as sole reliance on EF may overlook substantial hemodynamic impairment, underscoring its inadequacy as a standalone marker of systolic function. In this study, we complemented this clinical understanding with a technical investigation using a lumped-parameter cardiovascular model to simulate both healthy and pathological states, including MR. The model accurately reproduced hemodynamic parameters consistent with clinical benchmarks in healthy conditions and provided valuable insights into the effects of varying degrees of mitral valve leakage. By introducing a regurgitant pathway in the mitral valve representation, we quantified how progressive MR leads to reductions in FSV despite seemingly preserved LVEF — mirroring the clinical paradox observed in real-world patients. These simulation results reinforce the clinical observation that LVEF alone is an insufficient surrogate for cardiac performance in MR, particularly in moderate to severe cases. Instead, the forward stroke volume derived from left ventricular outflow tract velocity-time integral (LVOT VTI) offers a more physiologically relevant measure of true cardiac output.

## 5 Conclusion

This study presents a comprehensive evaluation of mitral regurgitation (MR) by combining clinical insights with a computational modeling approach. From a clinical perspective, MR is a prevalent and increasingly significant cardiovascular disorder that imposes a progressive burden on left ventricular function. It can obscure the detection of true cardiac dysfunction due to the limitations of conventional markers such as left ventricular ejection fraction (LVEF). Forward stroke volume (FSV) has emerged as a more physiologically accurate measure of effective cardiac output, particularly in moderate to severe MR, where total stroke volume and LVEF can remain deceptively preserved despite significant hemodynamic compromise. To explore this phenomenon quantitatively, we employed a closed-loop lumped-parameter model of the cardiovascular system, which was validated under healthy physiological conditions and subsequently adapted to simulate varying severities of MR. The simulation results clearly demonstrated how increasing mitral valve leakage progressively reduces FSV while leaving LVEF apparently unchanged, mirroring the clinical paradox observed in MR patients. These findings confirm the inadequacy of relying solely on LVEF for clinical decision-making in MR and support the incorporation of forward flow metrics such as LVOT VTI in patient evaluation.

## Data Availability

All data produced in the present study are available upon reasonable request to the authors

## Declarations

### Ethical Approval

Not applicable.

### Consent to Participate

Not applicable.

### Consent to Publish

Not applicable.

### Data Availability Statement

Not applicable.

### Authors’ Contributions

1. Amirahmad Saeedi: Development, simulation and analysis of the mathematical model.

2. Mobina Ghajar: Clinical justification and literature review.

3. Sara Tarkiani: Clinical justification and literature review.

4. Amin Ramezani: Results analysis and discussion.

## Author Biographies

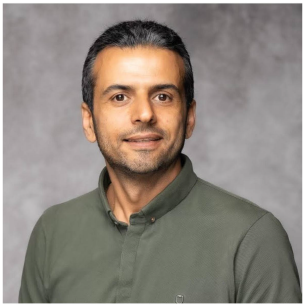

**Dr. Amin Ramezani**

Postdoctoral Fellow at the Division of Cardiac Surgery, Brigham and Women’s Hospital, Harvard Medical School, with a PhD in Artificial Intelligent Systems and Control. His research focuses on intelligent monitoring and control systems for complex dynamical systems, integrating Generative AI and Large Language Models (LLMs) into big data analytics for advancing healthcare. He specializes in deep learning-based medical image processing for early diagnosis and treatment, as well as the design and optimization of medical devices. His work is dedicated to preventative medicine and the development of predictive and prognostic tools, leveraging AI, computational modeling, and machine learning algorithms to enhance patient outcomes.

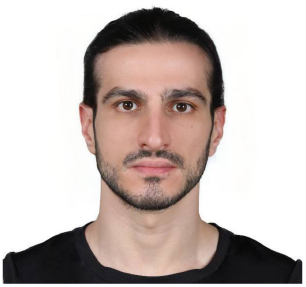

**Amirahmad Saeidi**

Amirahmad Saeidi received the B.S. degree in Electrical Engineering from Shahid Beheshti University, Iran, and the M.S. degree in Electrical Engineering (Control Systems) from Amirkabir University of Technology, Iran. He is currently researching the cardiovascular system.

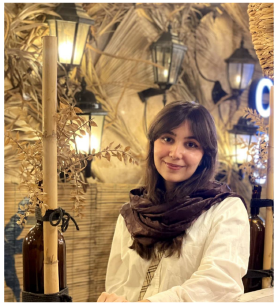

**Sara Tarkiani**

Final-year medical student at Zanjan University of Medical Sciences, with research in cardiovascular medicine focusing on arrhythmias, myocardial infarction and Artificial intelligence applications in cardiology—aspiring to advance cardiovascular knowledge through research and clinical care.

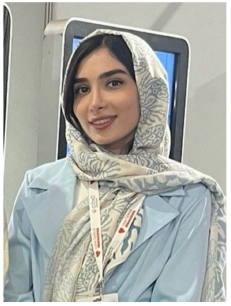

**Mobina Ghajar**

Final-year medical student at Zanjan University of Medical Sciences, with research experience in cardiovascular diseases, arrhythmias, and AI applications in cardiology.

